# Spillover Effects on Infant Caregiving and Domestic Work During the Poriborton Clean Cookstove Trial

**DOI:** 10.64898/2025.12.16.25342420

**Authors:** Md. Khobair Hossain, Neeloy Ashraful Alam, Elizabeth K Kirkwood, Sajia Islam, Sk Masum Billah, Md. Rashidul Azad, S M Rokonuzzaman, Camille Raynes-Greenow

**Affiliations:** International Centre for Diarrheal Disease Research, Bangladesh (icddr,b); University of Sydney, Sydney School of Public Health, Sydney, Camperdown, 2000, NSW Australia; School of Health Sciences, Queen’s Medical Centre, the University of Nottingham, United Kingdom; House of Research and Development (HRD), 56/9, Kalabagan, Dhaka, Dhaka 1205 Arnonld School of Public Health, University of South Carolina, 921 Assembly St, Columbia, SC 29208, United States

**Keywords:** Child Care, Infant Caregiving, Cookstoves, Qualitative methods, Time use, Household Work, Bangladesh

## Abstract

Quality care during early childhood is essential for life-long health, well-being, and productivity. Recognizing the global importance of advancing early childhood development strategies, this qualitative study conducted in-depth, face-to-face interviews with women, their husbands, and mothers-in-law to explore perceived shifts in childcare roles among participants of Poriborton Trial, a randomized controlled trial of Liquefied Petroleum Gas (LPG) cookstoves. These interviews aimed to capture diverse perspectives on evolving caregiving dynamics and explored mothers’ perspectives. Qualitative data were systematically analyzed using thematic analysis following an iterative process of coding and theme development.

We found that LPG stoves facilitated notable changes in caregiving. Firstly, LPG adoption reduced cooking time and decreased workload, which increased women’s time availability for childcare. Secondly, mothers reported a greater capacity to balance household tasks and childcare, fostering more frequent interaction with their children. Thirdly, the study identified enhanced childcare practices, with LPG use enabling women to be more responsive to their children’s needs particularly during illness and to maintain closer physical proximity.

The adoption of LPG stoves has multiple positive impacts on the childcare roles of breastfeeding women, empowering them to engage more fully in their children’s upbringing. Building on these qualitative insights into how LPG use may enhance childcare, future quantitative research is recommended to rigorously assess outcomes such as reductions in maternal stress, increased time availability for mothers, and changes in mother-child attachment attributable to LPG stove adoption.

## Background

In many low- and middle-income countries (LMICs), traditional gender roles remain deeply entrenched, with women primarily responsible for infant care and domestic work (1). Care provided during the first years of life is critical for child development, influencing physical health, cognitive growth, and emotional well-being (2–4). Quality early care during breastfeeding and early childhood can shape lifelong outcomes. However, these intensive caregiving phases coincide with significant demands on women’s time and energy, often creating a challenging environment for mothers (5,6).

In rural LMIC settings such as Bangladesh, new mothers experience challenges that are compounded by limited access to clean energy (7). Most households rely on biomass fuels such as wood, crop residues, and dung for cooking. While these fuels are inexpensive and locally available, their use imposes a heavy burden on women. The process of collecting, cutting, drying, and storing biomass fuel is labor-intensive and time-consuming, often requiring hours of physical effort each week (6). Cooking with biomass fuel also involves prolonged exposure to smoke and heat, which not only affects respiratory health but also contributes to fatigue and stress (8). These cumulative demands reduce the time and energy women can devote to childcare, potentially compromising the quality of care provided to infants (9,10).

The Poriborton trial was a community-based randomized controlled trial conducted in Sherpur district, Bangladesh, designed to evaluate the impact of transitioning from traditional biomass cooking to liquefied petroleum gas (LPG) on perinatal morbidity and mortality (11). As part of the intervention, participating households received LPG stoves and a continuous supply of LPG fuel during pregnancy until childbirth (11). While the primary focus of the trial was maternal and neonatal health outcomes, the intervention introduced a shift in household energy practices, raising the possibility of broader changes in child care roles.

Cooking with LPG offers several advantages over biomass fuel. It reduces cooking time, eliminates the need for fuel collection, and minimizes exposure to harmful smoke (7). These changes can substantially decrease women’s physical workload and stress, freeing time for other activities. Time savings and reduced fatigue could positively influence caregiving practices, enabling mothers to engage more actively with their children, respond promptly to their needs, and maintain closer emotional and physical contact. Improved caregiving during infancy is associated with better health, nutrition, and developmental outcomes, making this an important area of inquiry (3).

Evidence from other LMIC contexts suggests that clean cooking interventions can have unintended or spillover effects beyond health, including improvements in gender equity, household dynamics, and psychosocial well-being (12–14). However, these dimensions remain underexplored in Bangladesh, where cultural norms and economic constraints continue to shape women’s caregiving roles including domestic responsibilities. Understanding whether and how LPG adoption influences caregiving behaviors can provide important insights for designing interventions that address both health and social determinants of well-being.

This study therefore aims to explore the experiences of breastfeeding mothers who participated in the Poriborton trial. Specifically, it examines how the adoption of LPG stoves affected their ability to care for infants and manage domestic responsibilities. By focusing on caregiving practices, time allocation, and perceived stress, the study seeks to identify spillover effects of clean cooking interventions.

## Materials and Method

This study used a qualitative design to understand the unintended or spillover effects of using LPG for cooking, we followed a step-by-step process. First, we explored how using LPG cookstoves immediately influenced women’s experiences, and then we identified perceived changes in their roles and responsibilities. Next, we listed any changes to practices of the women due to cleaner cooking. Finally, we confirmed in follow-up interviews whether these changes were a result of using the LPG stove in the Poriborton trial.

### Participants

We recruited women who were currently participating in the intervention arm of Poriborton and their family members: husbands and their mothers-in-law who they reside with. Eligible women were those who were breastfeeding mothers from the intervention arm (12).

Participants were selected through purposive sampling based on having an infant younger than 24 months.

### Data Collection and analysis

We conducted in-depth interviews with participants in their households. We developed open-ended questions to allow for an in-depth exploration of the participants’ experiences and perceptions of the effect of LPG stove use on their childcare role. We used a team of six female and male qualitative researchers collected the data in November 2022. We conducted interviews in Bangla, and audio recorded them. We then transcribed the interviews verbatim into Bengali using the transcription software known as ‘TRANSKRIPTOR, and this data was imported into atlas.ti, a qualitative data analysis software for coding.

We used thematic analysis techniques, such as grouping comments and identifying patterns, resulting in categories and themes. Two researchers independently coded the data and discussed their codes to resolve any discrepancies. The codes were then grouped into categories to develop themes, which were reviewed and refined by the research team. Data were managed using atlas.ti software and backup copies were kept in password-protected computers accessed by the research team members only.

### Ethical Considerations

This study was approved by the ethical review committee of icddr,b (PR-17103) and the Human Research Ethics Committee of the University of Sydney, Australia (0/2018/HE000717). Informed written consent was obtained from all participants before the interviews.

## Results

The study included 21 women and 10 men. Among the women, 11 were breastfeeding mothers enrolled in the PORIBORTON trial, and ten were their mothers-in-laws. The rest ten male participants were the husbands of breastfeeding mothers.

There central themes emerged; resource enhancement for childcare through increased time management efficiency, enhanced Caregiver Capacity through reduced fatigue and stress, and Direct Improvement in Childcare Practices through increased opportunity for childcare (see table 1).

**Table 1:** Thematic Framework of Childcare Changes Linked to LPG Stove Use.

| (A) Code | (B) Category | (C) Theme |
| --- | --- | --- |
| Reduced cooking time<br>Increased parallel activities<br>Reduced cooking workload | Time Availability for Care<br>Streamlined Household Tasks<br>Reduced Domestic Burden | Theme 1: Resource Enhancement for Childcare through increased time management efficiency |
| Decrease in physical exhaustion<br>Decrease in mental stress of household roles | Improved Physical Well-being<br>Improved mental Well-being | Theme 2: Enhanced Caregiver Capacity through reduced fatigue and stress |
| Increase in mother-child attachment<br>Increase in caregiving during child illness<br>Increase in night-time care<br>Increase in quick responding to childcare needs | Enhanced Mother-Child Interaction<br>Increased Responsiveness<br>Proactive Caregiving | Theme 3: Direct Improvement in Childcare Practices through increased opportunity for childcare |

### Theme 1: Resource Enhancement for Childcare through increased time management efficiency

Mother participants who were primary cooks reported increased available time which was primarily due to the efficiency of LPG stove. They consistently reported that LPG stoves substantially reduced cooking time, lessened their cooking workload, and created opportunities for engaging in parallel activities. These direct benefits translated into several key changes in their daily routines: increased time availability specifically for childcare, the ability to streamline other household tasks, and a subsequent reduction in their overall domestic burden. Consequently, the adoption of LPG technology appears to provide mothers with ‘time’, a crucial resource which they can potentially allocate towards childcare responsibilities.

Every mother unanimously affirmed that this time-saving benefit has freed up time to engage more with their children. A mother described how she was able to take better care of her child after being an LPG stove user.

> *“Normally, I have to do all the housework and bring up the children. Most of the time it gets difficult to take care of my child. But now after using the LPG stove I save time and I get some (more) time to spend with my child.”* IDI-04

Another mother explained how the LPG stove reduced her cooking time by saying,

> *“Now LPG use has saved my time…and I have the time for feeding my baby myself and taking leisurely walks with her. I can even enjoy precious moments of play with her.”* IDI-07

A mother-in-law shared how her daughter-in-law has effectively saved time managing household chores through the use of the LPG stove.

> *“Before using LPG, my daughter-in-law was constantly rushed, spending hours on cooking.…Now, she has more time to care for her child, even holding them close whenever they cry.”* IDI-17

Several husbands shared that since their wives began using gas stoves, they could now complete their morning cooking and other tasks more time efficiently. One husband expressed,

> *“…Using LPG has helped my wife prepare meals and even reheat food in the morning. Not only does she have time to bathe her baby and even hold her when she has time[…] and it feels good that she can take care of the baby along with all her chores.”* IDI-27

Participants expressed their ability to engage in parallel activities while cooking. These included simultaneously cutting vegetables while boiling water, attending to other household chores, or even pursuing personal interests. Both mothers-in-law and husbands observed these changes and the adoption of parallel activities, and included women taking baths, sweeping the yard, cleaning rooms tending to cattle, all while food was cooking on the LPG stove.

> *“When I used to cook in clay stove, I had to stay with that one task for a long time, as a result of which the time goes by and the work does not progress. Now I can breastfeed my younger child, and also can sweep the rooms, leaving the food to cook on gas stove. During cooking now, I can perform additional 2-3 tasks simultaneously.”* IDI-10

A mother-in-law shared her observations about the multitasking abilities her daughter-in-law demonstrated while using the LPG stove.

> *“My daughter in law had to allocate much time in front of the clay stove when she used to cook using dry leaves. She had to complete her daily tasks one by one since she could not leave the cooking place. But now gas stove has brought about a change, she sweeps the yard or takes a bath when she cooks.”* IDI-20

Women and mothers-in-law alike noted that these parallel activities included childcare responsibilities and various other household tasks.

### Theme 2: Enhanced Caregiver Capacity through reduced fatigue and stress

The second prominent theme emerging from the data highlights an enhanced caregiver capacity among mothers using LPG stoves, attributed to a reported reduction in their fatigue and stress levels. Several mothers directly contrasted the labor-intensive nature of cooking with traditional clay stoves with the reduced physical burden experienced with LPG. Beyond the physical, mothers also expressed a belief that their mental stress related to household roles had decreased since adopting LPG, contributing to an overall improvement in their physical and mental well-being. This enhanced well-being was consistently linked to a greater ‘capacity’ for childcare. Husbands’ interviews often echoed this view, noting their wives’ increased capacity for childcare.

The qualitative data reveals a change in childcare practices among LPG-using mothers. They described increased opportunities to respond to their child, including during the night. In comparison to their experience of traditional stoves, LPG stoves allowed women to be more present and responsive to their children’s needs. These changes were recognized by all participants.

Moerover, mothers-in-law and husbands also observed a heightened level of emotional involvement of mothers in childcare.

#### Increasing care opportunities

Mothers reported being able to hold the child while cooking and this proved to be a new opportunity to take care of their child. One of the mothers shared how using the LPG stove enabled her to care for her infant while cooking.

> *“…My eldest baby wouldn’t stop crying if I didn’t cradle her in my arms. Unfortunately, I couldn’t always hold her close because our old clay stove required frequent refueling, filling the kitchen with fumes. My youngest baby’s fate is better because we have a gas stove and cooking has become more manageable. Now I can hold my younger child on my lap while preparing meals.*

> *It’s a remarkable feeling, as I couldn’t provide the same comfort to my elder child in the past.”* IDI-13

Another mother described how she managed to put her baby to sleep, while cooking continued.

> *“Every morning at 11 a.m., like clockwork, the baby goes to sleep once more. I have discovered that lying down and breastfeeding her during this time helps her fall asleep faster and more peacefully. Previously, I could not leave the pots unattended for fear of burning, which often caused delays in putting the baby to bed, and the baby would cry in discomfort. Now I have a gas stove, I can control the cooking process. I reduce the flame and can now take some time to sleep beside the baby, and she seems content and calm without any insistence on her part.”* IDI-16

Several mothers mentioned that breastfeeding became more manageable as mothers could save some time. The use of LPG stoves also reduced the likelihood of ignoring breastfeeding due to mother’s constant engagement with household work. Mothers believed that the convenience of LPG stoves enabled them to breast feed more frequently.

Few mothers-in-law noted that before adopting the gas stove, when the baby cried during cooking, the mother had to interrupt her tasks to attend to the baby. However, now mothers can respond to their babies while using the gas stove. One of the mothers in laws quoted her observation by saying,

> *“Since receiving the gas stove, my daughter in law’s outlook has become more positive, and she responds to the child promptly. She engages in conversation, sings, and interacts with my grandchild.”* IDI-29

### Theme 3: Direct Improvement in Childcare Practices through increased opportunity for childcare

The third theme emphasizes the reduced fatigue and stress experienced by participants as a result of using gas stoves. Several mothers directly contrasted the labor-intensive nature of cooking with traditional stoves with the reduced physical burden experienced with LPG. Beyond the physical, mothers also expressed a belief that their mental stress related to household roles had decreased since adopting LPG, contributing to an overall improvement in their physical and mental well-being. This enhanced well-being was consistently linked to a greater ‘capacity’ for childcare. Supporting this perspective, interviews with husbands frequently supported their wives’ increased ability to care for their children.

#### Decrease in physical exhaustion

Mothers expressed feeling less tired, less weak, and experiencing fewer bouts of dizziness after transitioning to LPG stoves. The reduced fuel collection, quicker cooking times and reduced physical exertion in the kitchen (from scrubbing blackened pots) contributed to their improved well-being.

> *“I live with my mother-in-law and my sister in-law, but I do most of the household works. I used to collect dry leaves outside, slash solid fuels. Further, I spent long hours cooking on clay stove which often left me fatigued and sometimes even dizzy due to sitting longer in front of the stove. After starting cooking in gas stove, I do not have to sit in front of the stove at a stretch. Now I use gas stove and I do not have to go outside to collect dry leaves or solid fuel which has greatly eased my physical burden. Now I get less tired and feel better physically.”* IDI-19

Moreover, many husbands and mothers-in-law reported that mothers displayed greater enthusiasm in caring for their children. A husband described the transformation in his wife resulting from a reduced workload, saying,

> *“My wife often used to have a serious mood in the afternoon, and I could understand with her hard work throughout the day. It is good that now she uses a gas stove, many of her demanding tasks are no longer necessary. When I return home in the afternoon, I find her in a cheerful mood.”* IDI-15

#### Decrease in mental stress of household roles

Mothers shared how LPG gas stoves allowed them to overcome the morning rush and worries about cooking describing a reduction in mental stress in that busy morning time. Women reported no longer feeling guilty over not being able to cook breakfast timely due to fuel shortages. A mother mentioned how her cooking related stress has decreased with the adoption of LPG stove.

> *“As a housewife I have to worry every time in my in-laws’ home wondering if I can cook quickly, provide breakfast for my children and above all, my husband. All these thoughts create mental stress for me. Now after being a gas stove user, I can ensure that my children can have breakfast before they go to school and my husband can start his day with a meal before going to work. So now my mental stress is eased.”* IDI-25

Several mothers-in-law noted the morning cooking stress experienced by mothers, who often grappled with the dilemma of collecting fuels or preparing for cooking. They underlined that the consistent availability of gas had notably eased the situation for mothers and relieved their morning cooking anxieties. A few husbands also remarked that their wives now seemed more at ease in the mornings compared to before.

A mother-in-law described her observations regarding the changes in timing and consistency of morning meal preparation by her daughter-in-law.

> *My daughter-in-law always tries to prepare breakfast for the whole family. But it was often difficult when she had to cook using dry leaves and straw. Sometimes there was no fuel, and she had to go out to collect it. Now, with the availability of LPG stoves, she prepares breakfast on time every day.* IDI-26

A husband noted that before using the LPG stove, his wife used to be exhausted from her daily workload. He further shared that she has now become able to care for their baby even during the night.

> *“My wife used to be exhausted from her full day’s work and would sometimes sleep through the night, even if the child cried. Since she started using a gas stove, she appears more attentive and capable of caring for the child, even during nighttime hours.”* IDI-33

A husband shared his opinion about his wife’s growing opportunities and improved efficiency in providing childcare after becoming an LPG stove user.

> *“…Using LPG has helped my wife prepare meals and even reheat food in the morning. Not only does she have time to bathe her baby and even hold her when she has time[…] and it feels good that she can take care of the baby along with all her chores.”* IDI-30

## Discussion

This study explores women’s experiences and changes to their caregiving roles after receiving an LPG stove and fuel due to participating in the Poriborton trial in rural Bangladesh. Overall, the impact on their time due to using LPG for cooking has been the key factor that led to changes in their caregiving practices, including child responsiveness, through a decrease in domestic workload burden attributable to household cooking, that has led to an increase in their well-being. This was experienced by the women and recognized by their immediate family, their husbands and their mothers-in-law. These findings indicate that adopting clean cooking may have benefits beyond clean household air and enable more child-responsive care of infants, through the pathway of increased time availability.

In prior research the time burden of using traditional stoves is well-established (14). Similarly, changes to time use using cleaner fuel for cooking has been demonstrated in previous research (15–16). In our study, in contrast to women’s experience with biomass fuel, LPG for cooking offered a consistent flame and increased heat, resulting in faster cooking times, that required less attention during cooking. These factors allowed the women, to conduct parallel activities, and thus save time, and therefore be more attentive to her child. A study showed that women who multitask during cooking can reduce their overall cooking time to some extent (17). The adoption of LPG stoves freed up time previously spent on the demanding tasks of biomass cooking, which participants reported allowed more frequent interaction with their children (18).

As trial participants, households were provided with free LPG fuel during the trial for a period of approximately seven months, which reduced the need to collect and prepare solid fuels, again ‘freeing’ up time. In a systematic review of time saving associated with adopting cleaner fuels, although there were only two included studies, the time spent collecting fuel was reduced between 1.2 to 1.5 hours daily, and cooking time was 0.7 fewer hours a day (16). Although the main focus of that review was the potential for income generation, our study who included breast feeding mothers, all used this extra time to provide more infant care, including breastfeeding. Breastfeeding is also a time-consuming activity, especially exclusive breastfeeding (19).

Our research is aligned with previous research that emphasized the importance of effective time management for breastfeeding mothers in providing better care for their children (20). Our study found that these women reduced their daily cooking domestic activities, and were able to engage in parallel activities, such as being more attentive to their children.

The increased availability as a caregiver associated with LPG stove use may have been through the mechanism of a reduction in fatigue and stress associated with the adoption of LPG stoves. Our results indicate a reported decrease in physical exhaustion and a reported lessening of mental stress related to their domestic roles. The process of collecting and preparing biomass or solid fuel, alongside other household duties, likely contributed to the physical strain experienced by these women.

Existing research has documented the physical demands of rural women’s responsibilities (21,22), suggesting that prolonged labor-intensive work can contribute to physical fatigue and potentially impact productivity and stress (14). The reported alleviation of physical exhaustion, as indicated by our participants, may have a positive influence on their childcare ability. Consistent with findings from a quantitative study suggesting that LPG use reduces the labor-intensive aspects of cooking (15), our research corresponds with a reported decrease in feelings of fatigue among women, which could potentially enhance their capacity to provide essential care to their children.

This, in turn, appeared to provide mothers with more time for rest and interaction with their children, potentially expanding their caregiving opportunities. This aligns with the findings of (24), who highlighted that effective parenting requires sustained physical and mental wellbeing. For women whose cooking workload caused exhaustion a decrease in that workload may have enabled them to conduct more parenting.

Several studies support our observations by highlighting the potential positive impact of LPG stove usage on women’s overall well-being, particularly in relation to household responsibilities (25,26, 13). This research points to a potential role of LPG gas stoves in mitigating the reported fatigue and mental stress experienced by breastfeeding women, which may, in turn, support their ability to provide quality care to their children. Consistent with studies in India (27–29), the time saved through LPG use appeared to provide mothers with more flexibility to attend to their children. This suggests a potential reallocation of women’s time within the realm of domestic responsibilities, allowing for a greater focus on responsive parenting.

### Strengths and Limitations

A key strength of this study lies in its triangulation of findings through interviews with mothers, their husbands, and mothers-in-law. This multi-perspective approach facilitated a richer understanding of changes in practices following the adoption of LPG and helped validate mothers’ accounts by incorporating the viewpoints of their spouses and mothers-in-law. A potential limitation is responder bias. Participants received free LPG stoves from the project team and knew the researchers were part of the project team, which may have positively influenced their interview responses. Further we have not been able to objectively assess parenting, breastfeeding or time-use and hence.

### Conclusion

The spill-over effect of participating in a LPG cookstove trial adds additional benefits for the participants. The LPG stoves reducing time spent cooking, and reduced the cookstove management, and fuel preparation associated with biomass cooking that enabled women to be more responsive to their infants. Participants also reported decreased fatigue and stress. These learnings suggest the potential of LPG stoves to alleviate the cooking burden and enhance childcare. This evidence can inform future programs that integrate energy access with maternal and child health strategies, contributing to broader goals of women empowerment and sustainable development.

Investing in clean cooking may improve parental well-being and improve parent responsiveness to their child. While this study provided initial insights into potential pathways of improved childcare associated with LPG adoption, future quantitative research could further investigate these relationships. Specifically, quantitative studies are recommended to measure the extent of reduction in mental stress, the quantifiable increase in mothers’ time availability, and the degree of change in mother-child attachment following LPG stove adoption.

## Data Availability

All files are available from the cloud server of the University of Sydney

## Acknowledgement

We gratefully acknowledge all participants for their invaluable contributions to this study. This research was supported by the National Health and Medical Research Council (Grant No. GNT2001264). We extend our sincere gratitude to the participants who generously shared their experiences as users of gas stoves. Their insights added major depth and richness to our findings.

Our heartfelt thanks go to the dedicated field researchers Md. Hafizur Rahman, Shayla Jesmin Nimmy, and Sultan Rohan whose tireless efforts in capturing the involved details of caregivers’ activities were important in ensuring the accuracy and reliability of the data collected.

## References

1. Kodali PB. Achieving Universal Health Coverage in Low- and Middle-Income Countries: Challenges for Policy Post-Pandemic and Beyond. Risk Manag Healthc Policy. 2023 Apr 6;16:607–621.

2. Richter L, Black M, Britto P, et al. Early childhood development: An imperative for action and measurement at scale. BMJ Glob Health. 2019;4:i154–i160.

3. Nelson CA, Scott RD, Bhutta ZA, Harris NB, Danese A, Samara M. Adversity in childhood is linked to mental and physical health throughout life. BMJ. 2020 Oct 28;371:m3048.

4. Clarke T. Children’s wellbeing and their academic achievement: The dangerous discourse of ‘trade-offs’ in education. Theory Res Educ. 2020;18(3):263–94.

5. Temple Newhook J, Newhook LA, Midodzi WK, Murphy Goodridge J, Burrage L, Gill N, et al. Poverty and breastfeeding: Comparing determinants of early breastfeeding cessation incidence in socioeconomically marginalized and privileged populations in the FiNaL study. Health Equity. 2017 Jun 1;1(1):96–102.

6. Rahman MA, Khan MN, Akter S, Rahman A, Alam MM, Khan MA, et al. Determinants of exclusive breastfeeding practice in Bangladesh: Evidence from nationally representative survey data. PLoS One. 2020 Jul 15;15(7):e0236080.

7. Haq I, Khan M, Chakma S, Hossain MI, Sarkar S, Rejvi MRA, Salauddin M, Sarker MMR. Determinants of household adoption of clean energy with its rural-urban disparities in Bangladesh. Sci Rep. 2024 Jan 29;14(1):2356.

8. Jessel S, Sawyer S, Hernández D. Energy, poverty, and health in climate change: a comprehensive review of an emerging literature. Front Public Health. 2019;7:357.

9. Chillrud SN, Ae-Ngibise KA, Gould CF, Owusu-Agyei S, Mujtaba M, Manu G, Burkart K, Kinney PL, Quinn A, Jack DW, Asante KP. The effect of clean cooking interventions on mother and child personal exposure to air pollution: results from the Ghana Randomized Air Pollution and Health Study (GRAPHS). J Expo Sci Environ Epidemiol. 2021 Jul;31(4):683–698.

10. Krishnapriya PP, Chandrasekaran M, Jeuland M, Pattanayak SK. Do improved cookstoves save time and improve gender outcomes? Evidence from six developing countries. Energy Econ. 2021 Oct;102:105456.

11. Ho EW, Strohmeier-Breuning S, Rossanese M, Charron D, Pennise D, Graham JP. Diverse health, gender and economic impacts from domestic transport of water and solid fuel: a systematic review. Int J Environ Res Public Health. 2021;18(19):[about 1 p.].

12. Raynes-Greenow C, Billah SM, Islam S, Rokonuzzaman SM, Tofail F, Kirkwood EK, et al. Reducing household air pollution exposure to improve early child growth and development; a randomized control trial protocol for the “Poriborton-Extension: The CHANge trial”. Trials. 2022 Jun 16;23(1):505.

13. Gould CF, Urpelainen J. LPG as a clean cooking fuel: adoption, use, and impact in rural India. Energy Policy. 2018;122:395–408.

14. Williams KN, Kephart JL, Fandino-Del-Rio M, Simkovich SM, Koehler K, Harvey SA, et al. Exploring the impact of a liquefied petroleum gas intervention on time use in rural Peru: a mixed methods study on perceptions, use, and implications of time savings. Environ Int. 2020;145:105932.

15. Simkovich SM, Williams KN, Pollard S, Dowdy D, Sinharoy S, Clasen TF, Puzzolo E, Checkley W. A systematic review to evaluate the association between clean cooking technologies and time use in low- and middle-income countries. Int J Environ Res Public Health. 2019;16(13):2277.

16. Petrokofsky G, Harvey WJ, Petrokofsky L, Ochieng CA. The importance of time-saving as a factor in transitioning from woodfuel to modern cooking energy services: a systematic map. Forests. 2021;12(9):1149.

17. Abdulai MA, Afari-Asiedu S, Carrion D, Ae-Ngibise KA, Gyaase S, Mohammed M, et al. Experiences with the mass distribution of LPG stoves in rural communities of Ghana. Ecohealth. 2018;15(4):757–67.

18. Horwood C, Alfers L, Masango-Muzindutsi Z, Dobson R, Rollins N. A descriptive study to explore working conditions and childcare practices among informal women workers in KwaZulu-Natal, South Africa: identifying opportunities to support childcare for mothers in informal work. BMC Pediatr. 2019;19:382.

19. Purkiewicz A, Regin KJ, Mumtaz W, Pietrzak-Fiećko R. Breastfeeding: the multifaceted impact on child development and maternal well-being. Nutrients. 2025 Apr 11;17(8):1326.

20. Yoosefi Lebni J, Mohammadi Gharehghani MA, Soofizad G, Khosravi B, Ziapour A, Irandoost SF. Challenges and opportunities confronting female-headed households in Iran: a qualitative study. BMC Womens Health. 2020;20(1):183.

21. Seedat S, Rondon M. Women’s wellbeing and the burden of unpaid work. BMJ. 2021;374:n1972.

22. Dahlgren A, Kecklund G, Akerstedt T. Different levels of work-related stress and the effects on sleep, fatigue, and cortisol. Scand J Work Environ Health. 2005;31(4):277–85.

23. Zdun-Ryzewska A, Nadrowska N, Blazek M, Bialek K, Zach E, Krywda-Rybska D. Parent’s stress predictors during a child’s hospitalization. Int J Environ Res Public Health. 2021;18(22):[about 1 p.].

24. Nomaguchi K, Milkie MA. Parenthood and well-being: a decade in review. J Marriage Fam. 2020 Feb;82(1):198–223.

25. Cassidy J, Jones JD, Shaver PR. Contributions of attachment theory and research: a framework for future research, translation, and policy. Dev Psychopathol. 2013 Nov;25(4 Pt 2):1415-34.

26. Gould CF, Urpelainen J. The gendered nature of liquefied petroleum gas stove adoption and use in rural India. J Dev Stud. 2020;56(7):1309–29.

27. Gupta S, Sayer LC, Pearlman J. Educational and type of day differences in mothers’ time availability for child care and housework. J Marriage Fam. 2021;83(3):786–802.

28. Younger A, Alkon A, Harknett K, Kirby MA, Elon L, Lovvorn AE, et al; HAPIN investigators. Effects of a LPG stove and fuel intervention on adverse maternal outcomes: a multi-country randomized controlled trial conducted by the Household Air Pollution Intervention Network (HAPIN). Environ Int. 2023 Aug;178:108059.

29. Purkiewicz A, Regin KJ, Mumtaz W, Pietrzak-Fiećko R. Breastfeeding: The multifaceted impact on child development and maternal well-being. Nutrients. 2025 Apr 11;17(8):1326.

